# Community prevalence of SARS-CoV-2 virus in England during May 2020: REACT study

**DOI:** 10.1101/2020.07.10.20150524

**Authors:** REACT Study Investigators, Steven Riley, Kylie E. C. Ainslie, Oliver Eales, Benjamin Jeffrey, Caroline E. Walters, Christina Atchison, Peter J. Diggle, Deborah Ashby, Christl A. Donnelly, Graham Cooke, Wendy Barclay, Helen Ward, Graham Taylor, Ara Darzi, Paul Elliott

## Abstract

**Background:** England has experienced one of the highest rates of confirmed COVID-19 mortality in the world. SARS-CoV-2 virus has circulated in hospitals, care homes and the community since January 2020. Our current epidemiological knowledge is largely informed by clinical cases with far less understanding of community transmission.

**Methods:** The REal-time Assessment of Community Transmission (REACT) study is a nationally representative prevalence survey of SARS-CoV-2 virus swab-positivity in the community in England. We recruited participants regardless of symptom status.

**Results:** We found 159 positives from 120,610 swabs giving an average prevalence of 0.13% (95% CI: 0.11%,0.15%) from 1st May to 1st June 2020. We showed decreasing prevalence with a halving time of 8.6 (6.2, 13.6) days, implying an overall reproduction number R of 0.57 (0.45, 0.72). Adults aged 18 to 24 yrs had the highest swab-positivity rates, while those >64 yrs had the lowest. Of the 126 participants who tested positive with known symptom status in the week prior to their swab, 39 reported symptoms while 87 did not, giving an estimate that 69% (61%,76%) of people were symptom-free for the 7 days prior testing positive in our community sample. Symptoms strongly associated with swab-positivity were: nausea and/or vomiting, diarrhoea, blocked nose, loss of smell, loss of taste, headache, chills and severe fatigue. Recent contact with a known COVID-19 case was associated with odds of 24 (16, 38) for swab-positivity. Compared with non-key workers, odds of swab-positivity were 7.7 (2.4, 25) among care home (long-term care facilities) workers and 5.2 (2.9, 9.3) among health care workers. However, some of the excess risk associated with key worker status was explained by recent contact with COVID-19 cases. We found no strong evidence for geographical variability in positive swab results.

**Conclusion:** Our results provide a reliable baseline against which the impact of subsequent relaxation of lockdown can be assessed to inform future public health efforts to control transmission.

## Introduction

To date, the UK has experienced the third highest number of reported COVID-19 deaths after the United States and Brazil^1^. In response to rapid growth in health care demand, lockdown was implemented nationally on 23rd March and daily mortality in the UK peaked around 11th April. As of 9th July 2020, 44,517 deaths in people with a positive test had occurred in the UK^2^.

Until recently, testing for severe acute respiratory syndrome coronavirus 2 (SARS-CoV-2, cause of COVID-19) has focused mainly on health care^3,4^ and care home settings^5,6^. While there is evidence of transmission of SARS-CoV-2 virus and high levels of mortality in these settings, less attention has been paid to levels of the virus circulating in the community, especially among those not exhibiting symptoms. This is a critical knowledge gap since community transmission will play an increasingly important part in the epidemiology of the COVID-19 pandemic as lockdown measures are lifted^7^.

Understanding patterns of infection in the population and the risk by social, demographic and geographical factors is therefore crucial to help control the epidemic and prevent a second wave of infection. We report here findings from the REal-time Assessment of Community Transmission (REACT) study, a large population-based programme designed to establish prevalence of current SARS-CoV-2 infection across England toward the end of the lockdown period. In addition, it provides an opportunity to measure the growth rate of prevalence of infection in the community and hence infer the reproduction number (R).

## Methods

### Recruitment of participants

We sent a letter to a nationally representative sample of the population among the 315 lower-tier local authorities (LTLAs) in England. Participants were randomly selected from the

National Health Service (NHS) list of patients registered with a general practitioner, aged five yrs and over.

### Covariates

Data on age, sex, address and postcode were available from the NHS register. Information on key worker status, ethnicity, smoking, contact with known or suspected COVID-19 cases, and symptoms were obtained by online questionnaire or telephone. Participants could either report no symptoms or select one or more from a list of 26 symptoms in the past six months, and whether or not they had experienced symptoms in the last week. Among those who reported symptoms in the last week, we were not able to identify which of their symptoms in the previous 6 months had occurred in the previous week. For analysis, we assumed that any of the reported symptoms could have occurred in the last week.

### Nose and throat swab collection, sample handling and testing

Participants were given written and video instructions to obtain a self-administered nose and throat swab (parent /guardian administered for children aged 5-12 years). Initially swabs in viral transport medium were used and sent to one of four Public Health England (PHE) laboratories for analysis (n= 8,595 swabs with reported result). Subsequently we switched to obtaining dry swabs (n= 112,025). We asked participants to refrigerate the sample and book pick-up of the swab within 24 hours. Swabs were collected by courier and delivered to a commercial laboratory using a cold chain (4° to 8°C) to maintain sample integrity. Samples were tested using reverse-transcription--polymerase-chain-reaction (RT-PCR). A calibration study was undertaken to ensure comparability of test results across study laboratories and results for the commercial laboratory and PHE samples were then combined.

### PCR Calibration experiments

We observed that the proportion of positive results from the commercial laboratory was substantially higher than from the Public Health England (PHE) laboratories. It was apparent that the commercial laboratory was routinely reporting as positive, on testing by RT-PCR, samples with high Ct values for the N-gene target, although the E-gene target was not detected.

To reconcile these differences, we conducted three separate calibration experiments. First, 10 RNA extraction plates were sent from the commercial laboratory to two NHS accredited laboratories for blinded re-analysis. Results were concordant for 919 negative samples and all 40 controls. We detected viral RNA in 11 of the 19 samples reported positive by the commercial laboratory (N-gene Ct-value range 16.5 to 40.7); 10 of these 11 samples had an N-gene Ct value < 37. Second, the commercial laboratory conducted a serial dilution experiment of known positive samples with high viral load to assess Ct thresholds at the limit of detection. Third, a further 40 unblinded positive samples (on 19 plates) with Ct values (N-gene) > 35 (range 35.7 to 46.8) and without a signal for the E-gene were selectively re-analysed in a PHE reference laboratory; SARS CoV0-2 RNA was detected in 15/40 (38%) (2/4 with N-gene Ct value < 37). As a result of these calibration experiments, we report swab-positivity for positive samples reported by the commercial laboratory where N-gene Ct values < 37 or where virus was detected by both N-gene and E-gene targets.

### Analysis

To investigate association of different covariates with swab-positivity, we performed univariate logistic regression to obtain unadjusted odds ratios and 95% confidence intervals. Test result (positive/negative) was used as the outcome variable. We also used multi-variable models to adjust for age and sex and then additionally ethnicity, region, and key worker status. When assessing the difference in test result by age, we compared models with a categorical term for age (Table 1) as well as a smooth term.

**Table 1.** Unadjusted and adjusted odds ratios for logistic models of swab-positivity.

| Characteristic | Unadjusted odds ratio (95% CI) | Odds ratios adjusted for age and sex (95% CI) | Jointly adjusted odds ratios (95% CI) | Jointly adjusted odds ratios with smooth term for age (95% CI) |
| --- | --- | --- | --- | --- |
| Sex (n=120,620) |  |  |  |  |
| Male | reference | reference | reference | reference |
| Female | 0.99 ( 0.7 , 1.4 ) | 0.97 ( 0.71 , 1.3 ) | 0.84 ( 0.58 , 1.2 ) | 0.85 ( 0.59 , 1.2 ) |
| Age group (yrs, n=120,620) |  |  |  |  |
| 5 - 12 | 1.1 ( 0.61 , 2.1 ) | 1.1 ( 0.61 , 2.1 ) | 0.65 ( 0.22 , 1.9 ) | See Fig. 2 for smooth term |
| 13 - 17 | 1.1 ( 0.52 , 2.2 ) | 1.1 ( 0.52 , 2.2 ) | 1.6 ( 0.67 , 3.7 ) |  |
| 18 - 24 | 2.0 ( 1.1 , 3.6 ) | 2.0 ( 1.1 , 3.6 ) | 1.7 ( 0.84 , 3.6 ) |  |
| 25 - 34 | 1.2 ( 0.69 , 2.2 ) | 1.2 ( 0.69 , 2.2 ) | 1.3 ( 0.66 , 2.4 ) |  |
| 35 - 44 | reference | reference | reference |  |
| 45 - 54 | 0.97 ( 0.6 , 1.7 ) | 0.97 ( 0.56 , 1.7 ) | 1.0 ( 0.56 , 1.9 ) |  |
| 55 - 64 | 0.68 ( 0.4 , 1.2 ) | 0.68 ( 0.37 , 1.2 ) | 0.80 ( 0.42 , 1.5 ) |  |
| 65 + | 0.52 ( 0.3 , 0.97 ) | 0.52 ( 0.28 , 0.97 ) | 0.53 ( 0.24 , 1.2 ) |  |
| Region (n=120,620) |  |  |  |  |
| North East | 0.73 ( 0.29 , 1.9 ) | 0.73 ( 0.29 , 1.9 ) | 0.70 ( 0.25 , 2.0 ) | 0.70 ( 0.25 , 2.0 ) |
| North West | 1.0 ( 0.61 , 1.7 ) | 1.0 ( 0.61 , 1.7 ) | 1.1 ( 0.60 , 1.9 ) | 1.1 ( 0.61 , 1.9 ) |
| Yorkshire and The Humber | 0.60 ( 0.27 , 1.3 ) | 0.6 ( 0.27 , 1.3 ) | 0.43 ( 0.15 , 1.2 ) | 0.43 ( 0.15 , 1.2 ) |
| East Midlands | 0.77 ( 0.44 , 1.3 ) | 0.77 ( 0.44 , 1.3 ) | 0.70 ( 0.37 , 1.3 ) | 0.70 ( 0.37 , 1.3 ) |
| West Midlands | 1.1 ( 0.64 , 1.9 ) | 1.1 ( 0.64 , 1.9 ) | 1.1 ( 0.56 , 2.00 ) | 1.1 ( 0.56 , 2.00 ) |
| East of England | 0.68 ( 0.39 , 1.2 ) | 0.67 ( 0.38 , 1.2 ) | 0.77 ( 0.42 , 1.4 ) | 0.77 ( 0.42 , 1.4 ) |
| London | 1.3 ( 0.78 , 2.2 ) | 1.2 ( 0.73 , 2.1 ) | 0.98 ( 0.51 , 1.9 ) | 0.98 ( 0.51 , 1.9 ) |
| South East | reference | reference | reference | reference |
| South West | 0.37 ( 0.16 , 0.87 ) | 0.38 ( 0.16 , 0.89 ) | 0.39 ( 0.15 , 1.0 ) | 0.39 ( 0.15 , 1.0 ) |
| Key worker status (n=89,695) |  |  |  |  |
| Health care worker | 5.2 ( 2.9 , 9.3 ) | 5.5 ( 3.0 , 9.9 ) | 5.4 ( 3.0 , 9.9 ) | 5.2 ( 2.9 , 9.5 ) |
| Care home worker | 7.7 ( 2.4 , 25 ) | 8.4 ( 2.5 , 28 ) | 8.4 ( 2.5 , 28 ) | 8.3 ( 2.5 , 28 ) |
| Other key worker | 1.9 ( 1.1 , 3.1 ) | 1.8 ( 1.1 , 3.1 ) | 1.9 ( 1.1 , 3.2 ) | 1.9 ( 1.1 , 3.2 ) |
| Other worker | reference | reference | reference | reference |
| Not regular worker | 1.3 ( 0.8 , 2.0 ) | 1.5 ( 0.87 , 2.6 ) | 1.5 ( 0.86 , 2.6 ) | 1.6 ( 0.95 , 2.6 ) |
| Contact with COVID case (n=92,941) |  |  |  |  |
| No contact | reference | reference | -- | -- |
| Contact with confirmed case | 24 ( 16 , 38 ) | 23 ( 14 , 36 ) | -- | -- |
| Contact with suspected case | 5.3 ( 2.2 , 12 ) | 4.9 ( 2.0 , 12 ) | -- | -- |
| Ethnicity (n=120,620) |  |  |  |  |
| White | reference | reference | reference | reference |
| Asian | 1.9 ( 1.0 , 3.5 ) | 1.7 ( 0.92 , 3.2 ) | 1.7 ( 0.86 , 3.5 ) | 1.7 ( 0.86 , 3.5 ) |
| Black | 1.3 ( 0.3 , 5.4 ) | 1.3 ( 0.31 , 5.1 ) | 1.4 ( 0.34 , 5.8 ) | 1.4 ( 0.33 , 5.7 ) |
| Mixed | 1.4 ( 0.5 , 3.8 ) | 1.2 ( 0.42 , 3.1 ) | 0.9 ( 0.22 , 3.7 ) | 0.86 ( 0.21 , 3.5 ) |
| Other | 1.5 ( 0.4 , 6.0 ) | 1.4 ( 0.35 , 5.7 ) | 0.89 ( 0.12 , 6.4 ) | 0.88 ( 0.12 , 6.4 ) |
| Smoking status (n=22,528) |  |  |  |  |
| Currently smoke cigarettes | 1.4 ( 0.4 , 4.6 ) | 1.2 ( 0.36 , 4.3 ) | -- | -- |
| Do not currently smoke cigarettes | reference | reference | -- | -- |

To investigate trends in swab positivity rate over time, we used an exponential decay model, assuming that the number of positive samples each day arose from a binomial distribution. Posterior credibility intervals were obtained using a bivariate Metropolis-Hasting algorithm with uniform priors. We used day of swabbing (reported for 72% of participants for whom a laboratory result was available) or, if unavailable, day of collection (increasing the number of participants for which a date was available to 92%). To estimate R^8^, we assumed a generation time distribution of six days, comprising exponential latent and infectious periods of three days each.

Clustering of prevalence at the level of LTLAs was assessed by testing the null hypothesis that the number of positive swabs from each LTLA arose from a binomial process with fixed probability q from n samples, where n was the number of tested swabs from that LTLA. We assessed our power to detect clustering in LTLAs by hypothesising that (1-q) LTLAs would have no swab-positive with the remaining infection being distributed evenly among the remaining proportion q of LTLAs. If that were the case, our study was powered to detect q < ∼0.6 (that is clustering in up to 60% of LTLAs) with 80% power (Figure 1).We present results of the main data using complete case analysis without imputation. Analysis was conducted in R version 4.0.0^9^. Logistic regression models were implemented in the base function glm or with smooth function using the mgcv package^10^. We used changes in Akaike information criterion (AIC) to assess improvement in model performance.

**Figure 1.**
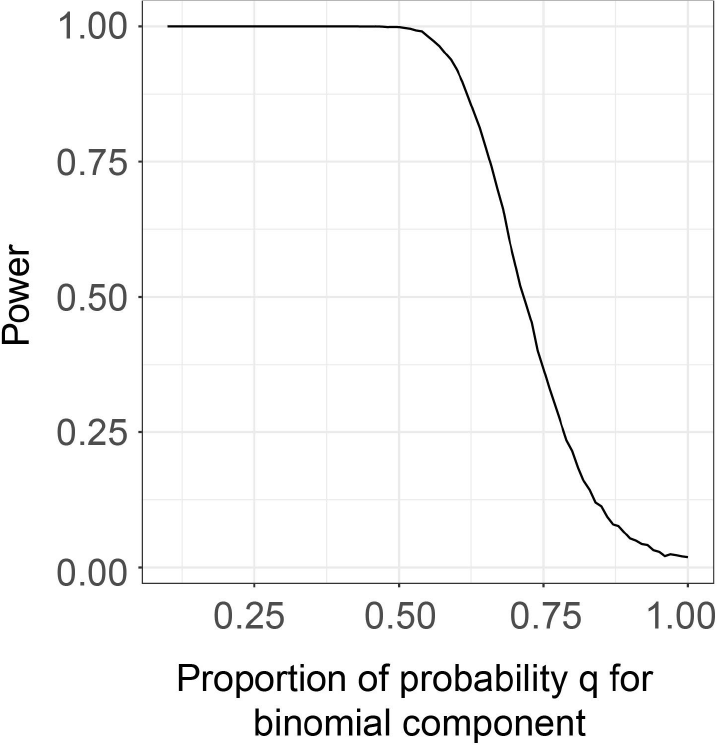
Power to detect clustering. Power of the chi-squared test to detect clustering at the LTLA level 0.05 significance for different degrees of clustering. Clustering is represented by q, the proportion of LTLA’s that follow a binomial distribution of positive results with the other 1-q LTLAs having fixed zero positive results (a zero-inflated distribution). 10,000 distributions among the LTLAs of the 159 positive samples are generated for each value of q considered. A chi-squared test is then completed for each distribution comparing the frequency of LTLA with each number of positive samples to that of the expected distribution assuming a purely binomial probability distribution. The power is defined as the proportion of chi-squared tests with a p-value less than 0.05.

### Ethics

We obtained research ethics approval from the South Central-Berkshire B Research Ethics Committee (IRAS ID: 283787).

## Results

### Sample and response rate

Letters were sent to 394,970 persons (base sample), of whom 161,771 (41%) (or parent/guardian) registered for the study, were sent a nose and throat swab kit, and were asked to complete a questionnaire. Swab results were available for 120,620 individuals (achieved sample, 31% of base sample). The achieved sample was broadly representative of the base sample although males, adults aged 18 to 34 yrs and London region were to an extent under-represented (Table 2).

**Table 2.** Characteristics of: all persons approached (n=394,970); those registered to whom a swab was sent (n=161,771); those for which a swab result was available (n=120,620); and those who were swab-positive for SARS-CoV-2 (n=159).

| Characteristic | All persons approached | All persons registered | All persons with swab result | SARS-CoV-2 positive |
| --- | --- | --- | --- | --- |
| Gender -- n (%) |  |  |  |  |
| Male | 196,732 (50) | 74,889 (46) | 55,064 (46) | 73 (46) |
| Female | 198,238 (50) | 86,882 (54) | 65,556 (54) | 86 (54) |
| Age group (yrs) -- n (%) |  |  |  |  |
| 5 - 12 | 39,268 (10) | 15,644 (10) | 10,573 (9) | 17 (11) |
| 13 - 17 | 23,098 (6) | 9,589 (6) | 7,293 (6) | 11(7) |
| 18 - 24 | 33,803 (9) | 10,370 (6) | 6,916 (6) | 19 (12) |
| 25 - 34 | 58,840 (15) | 19,861 (12) | 13,317 (11) | 23 (14) |
| 35 - 44 | 55,888 (14) | 22,754 (14) | 16,372 (24) | 23 (14) |
| 45 - 54 | 56,950 (14) | 27,215 (17) | 20,617 (17) | 28 (18) |
| 55 - 64 | 51,390 (13) | 26,288 (16) | 20,980 (17) | 20 (13) |
| 65 + | 75,733 (19) | 30,050 (19) | 24,552 (20) | 18 (11) |
| Region -- n(%) |  |  |  |  |
| North East | 15,324 (4) | 6,344 (4) | 4,511 (4) | 5 (3) |
| North West | 53,304 (13) | 21,590 (13) | 15,643 (13) | 24 (15) |
| Yorkshire and The Humber | 24,908 (6) | 10,628 (7) | 7,774 (6) | 7 (4) |
| East Midlands | 46,083 (12) | 20,146 (12) | 15,429 (13) | 18 (11) |
| West Midlands | 36,472 (9) | 15,016 (9) | 11,335 (9) | 19 (12) |
| East of England | 52,859 (13) | 23,189 (14) | 17,598 (15) | 18 (11) |
| London | 55,606 (14) | 15,247 (9) | 10,487 (9) | 21 (13) |
| South East | 79,671 (20) | 35,552 (22) | 27,142 (23) | 41 (26) |
| South West | 30,743 (8) | 14,059 (9) | 10,691 (9) | 6 (4) |

### Overall prevalence of swab-positivity

Out of 120,610 swab results, 159 were positive giving an overall unadjusted prevalence of 0.13% (0.11%,0.15%) for the period 1st May to 1st June 2020. This corresponds to an estimated 74,000 (63,000, 86,000) prevalent infections of SARS-CoV-2 virus in England on average which was unchanged when we adjusted for the age-sex distribution of the base sample.

### Decline in prevalence and estimate of reproduction number

We obtained strong evidence that prevalence was decreasing during the study period with a halving time of 8.6 (6.2, 14) days, corresponding to a decay rate of 0.081 (0.051, 0.11) per day (Figure 2). We estimated the reproduction number R to be 0.57 (0.45, 0.72).

**Figure 2.**
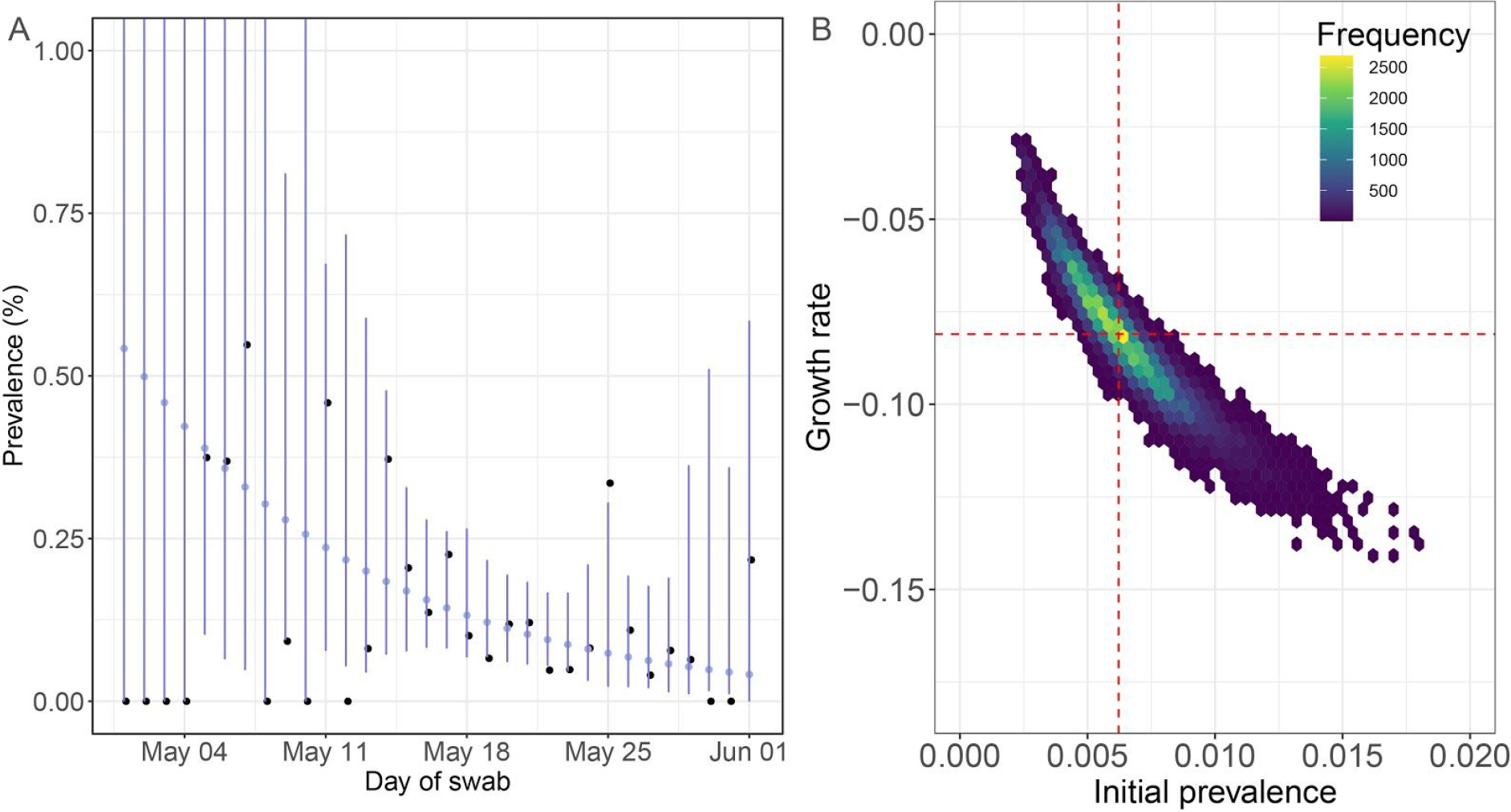
Temporal trends in swab positivity. **A** black points show daily observed prevalence by day of swab. Violet points show the best fit exponential decay line. Violet lines show binomial prediction intervals (95%) for the best fit. **B** bivariate posterior credibility interval for exponential decay model as a function of prevalence on 1st May (x-axis) and growth rate (y-axis).

We used a regression framework to test that the rate of decay did not vary substantially over the time of our study: a linear univariate model for day of swab as a covariate for swab-positivity had the same AIC as a univariate model with a smooth term for day of swab, with the smooth term producing a straight line (not shown).

### Trends with sex, ethnicity and age

Prevalence of swab positivity for males of 0.13% (0.11%,0.17%) was similar to that for females of 0.13% (0.11%, 0.16%) (Table 2). Asian participants (predominantly South Asian) were more likely to be swab-positive than white participants, with an unadjusted odds ratio of 1.9 (1.0, 3.5), although this reduced to 1.7 (0.86, 3.5) on adjustment (Table 1, Table 3). Adults aged 18 to 24 yrs had highest rates of swab positivity while those >64 yrs had lowest rates (Figure 3A, Table 4 in Supplement). A smooth term for age was better supported by the data than a categorical term in the adjusted model (Table 1, ΔAIC = 8) and its shape was largely unchanged when compared with a univariate model of age (also using a smooth term, Figure 3B). Unadjusted rates in children and young people aged 5 to 17 yrs were similar to those in adults aged 25 to 34, and remained so with a smooth term for age in the adjusted model.

**Table 3.** Prevalence by key characteristics.

| Characteristic | Persons with swab | SARS-CoV-2 positive | Prevalence (95% CI) |
| --- | --- | --- | --- |
| **Key worker status** |  |  |  |
| Health care worker | 3,801 | 18 | 0.47% ( 0.30% , 0.75% ) |
| Care home worker | 424 | 3 | 0.71% ( 0.24% , 2.06% ) |
| Other key worker | 16,364 | 28 | 0.17% ( 0.12% , 0.25% ) |
| Other worker | 32,588 | 30 | 0.09% ( 0.06% , 0.13% ) |
| Not regular worker | 36,518 | 42 | 0.12% ( 0.09% , 0.16% ) |
| Not known | 30,925 | 38 | 0.12% ( 0.09% , 0.17% ) |
| **Contact with COVID case** |  |  |  |
| No contact | 91,047 | 96 | 0.11% ( 0.09% , 0.13% ) |
| Contact with confirmed case | 1,000 | 25 | 2.50% ( 1.70% , 3.66% ) |
| Contact with suspected case | 894 | 5 | 0.56% ( 0.24% , 1.30% ) |
| Not known | 27,679 | 33 | 0.12% ( 0.08% , 0.17% ) |
| **Ethnicity** |  |  |  |
| White | 111,502 | 140 | 0.13% ( 0.11% , 0.15% ) |
| Asian | 4,578 | 11 | 0.24% ( 0.13% , 0.43% ) |
| Black | 1,188 | 2 | 0.17% ( 0.05% , 0.61% ) |
| Mixed | 2,281 | 4 | 0.18% ( 0.07% , 0.45% ) |
| Other | 1,071 | 2 | 0.19% ( 0.05% , 0.68% ) |
| **Smoking status** |  |  |  |
| Currently smoke cigarettes | 2,597 | 3 | 0.12% ( 0.04% , 0.34% ) |
| Do not currently smoke cigarettes | 19,931 | 17 | 0.09% ( 0.05% , 0.14% ) |
| Not known | 98,092 | 139 | 0.14% ( 0.12% , 0.17% ) |

**Table 4.** Prevalence by age.

| Age category | All participants | SARS-CoV-2 Positive | Prevalence % (95% CI) |
| --- | --- | --- | --- |
| 5 - 12 | 10,573 | 17 | 0.16% ( 0.10% , 0.26% ) |
| 13 - 17 | 7,293 | 11 | 0.15% ( 0.08% , 0.27% ) |
| 18 - 24 | 6,916 | 19 | 0.27% ( 0.18% , 0.43% ) |
| 25 - 34 | 13,317 | 23 | 0.17% ( 0.12% , 0.26% ) |
| 35- 44 | 16,372 | 23 | 0.14% ( 0.09% , 0.21% ) |
| 45 - 54 | 20,617 | 28 | 0.14% ( 0.09% , 0.20% ) |
| 55 - 64 | 20,980 | 20 | 0.10% ( 0.06% , 0.15% ) |
| 65 + | 24,552 | 18 | 0.07% ( 0.05% , 0.12% ) |

**Figure 3.**
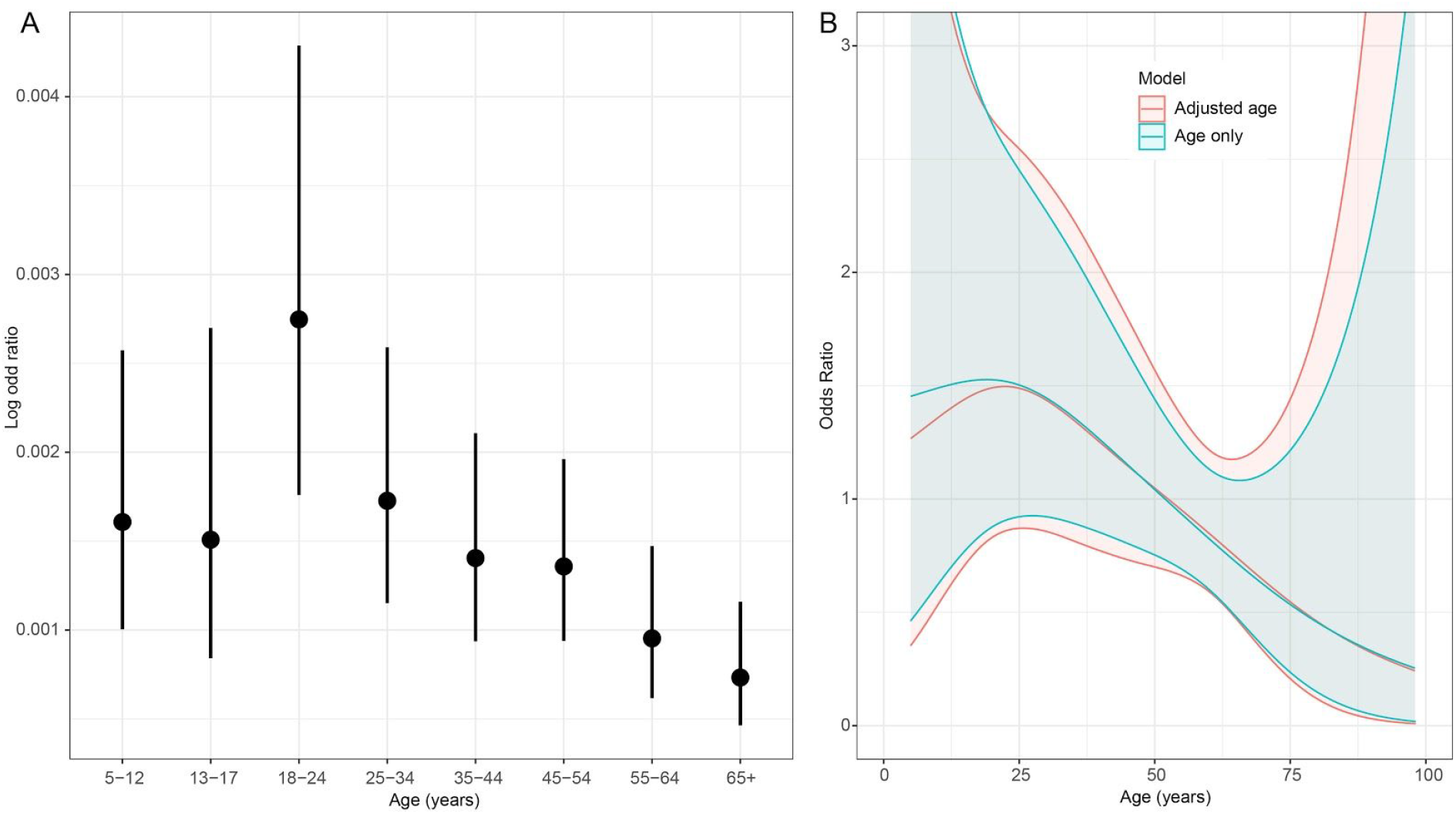
**A** Prevalence (%) by age and 95% CIs. **B** Curves from smooth terms in unadjusted (blue) and adjusted (red) logistic regression models of odds of swab-positivity. Regions are 95% CIs. See Table 1 for details of the adjusted model.

The trend in age was not explained by key worker status (see below). The adjusted model including a smooth term for age and key worker status was preferred to the model with a smooth age term alone (ΔAIC = 37) but the shape of the trend was conserved (Figure 3B).

### Contact with COVID-19 cases

Information was provided about possible contact with COVID-19 cases for 92,941 participants. Compared to a person who did not report contact with a COVID-19 case, those who reported contact with a confirmed case had odds of 24 (16, 38) of testing positive; odds were 5.3 (2.2, 12) for contact with a suspected case. These findings were essentially unchanged after adjustment for age and sex (Table 1).

### Key workers

The prevalence of infection was highest among care home workers, 0.71% (0.24%, 2.06%), followed by health care workers 0.47% (0.30%, 0.75%) and other key workers 0.17% (0.12%, 0.25%) (Table 3). Compared with non-key workers, unadjusted odds of testing positive were 7.7 (1.8, 22) in care home workers, 5.2 (2.8, 9.2) in health care workers and 1.8 (1.1, 3.1) in other key workers. Odds were similar or larger after adjustment for covariates not including contact with a COVID-19 case (Table 1). However, care home workers and health care workers were much more likely to report a recent contact with a COVID-19 case (Table 5), which contributed to the excess risk in those groups (not shown).

**Table 5.** Contact with COVID-19 case by key worker status.

| Key worker status | Contact with COVID-19 case |  |  |  |  |  |  |  | Total |
| --- | --- | --- | --- | --- | --- | --- | --- | --- | --- |
|  | No contact (%) |  | Contact with confirmed case |  | Contact with suspected case |  | Not known |  |  |
| Health care worker | 2,961 | 78% | 539 | 14% | 301 | 8% | 0 | 0% | 3,801 |
| Care home worker | 364 | 86% | 38 | 9% | 22 | 5% | 0 | 0% | 424 |
| Other key worker | 15,996 | 98% | 173 | 1% | 195 | 1% | 0 | 0% | 16,364 |
| Other worker | 32,339 | 99% | 89 | 0% | 160 | 0% | 0 | 0% | 32,588 |
| Not fulltime worker | 36,201 | 99% | 132 | 0% | 183 | 1% | 2 | 0% | 36,518 |
| Not known | 3,186 | 10% | 29 | 0% | 33 | 0% | 27,677 | 89% | 30,925 |

### Symptoms and swab-positivity

Of the 126 participants who tested positive with known symptom status in the week prior to their swab, 39 reported symptoms while 87 did not, giving an estimate that 69% (61%,76%) of people were symptom-free for the 7 days prior testing positive in our community sample. For those reporting symptoms in the last week (see Methods), the following were strongly associated with a positive swab result; nausea and/or vomiting, diarrhoea, blocked nose, loss of smell, loss of taste, headache, chills and severe fatigue (Table 6).

**Table 6.** Risk of positive swab for participants who reported symptoms in the last week (n=93,037). Boldface type highlights those symptoms with p<0.01.

| Symptoms<sup>1</sup> | All persons<br>REACT-1 | SARS-CoV-2<br>Positive | Odds Ratio | p-value |
| --- | --- | --- | --- | --- |
| Decrease in appetite | 2,123 | 7 | 2.76 (1.28, 5.97) | 0.01 |
| **Nausea and/or vomiting** | **2,531** | **9** | **2.98 (1.50, 5.92)** | **< 0.01** |
| **Diarrhoea** | **4,467** | **13** | **2.44 (1.36, 4.37)** | **< 0.01** |
| Abdominal pain/tummy ache | 4,669 | 10 | 1.79 (0.93, 3.45) | 0.08 |
| Runny nose | 9,610 | 20 | 1.74 (1.07, 2.83) | 0.03 |
| Sneezing | 9,759 | 21 | 1.80 (1.12, 2.90) | 0.02 |
| **Blocked nose** | **7,518** | **20** | **2.23 (1.37, 3.62)** | **< 0.01** |
| Sore eyes | 5,634 | 10 | 1.48 (0.77, 2.86) | 0.24 |
| **Loss of sense of smell** | **1,421** | **12** | **7.11 (3.88, 13.02)** | **< 0.01** |
| **Loss of sense of taste** | **1,436** | **11** | **6.44 (3.43, 12.09)** | **< 0.01** |
| Sore throat | 8,170 | 15 | 1.53 (0.89, 2.66) | 0.13 |
| Hoarse voice | 2,799 | 5 | 1.49 (0.61, 3.68) | 0.38 |
| **Headache** | **10,369** | **24** | **1.94 (1.23, 3.68)** | **< 0.01** |
| Dizziness | 4,040 | 8 | 1.66 (0.80, 3.42) | 0.17 |
| Shortness of breath affecting normal activities | 3,275 | 5 | 1.28 (0.52, 3.14) | 0.60 |
| New persistent cough | 2,929 | 9 | 2.57 (1.29, 5.11) | 0.01 |
| Tightness in chest | 3,873 | 9 | 1.94 (0.98, 3.86) | 0.06 |
| Chest pain | 2,226 | 5 | 1.88 (0.76, 4.63) | 0.17 |
| Fever (feeling too hot) | 3,045 | 8 | 2.20 (1.06, 4.54) | 0.03 |
| **Chills (feeling too cold)** | **3,377** | **11** | **2.73 (1.45, 5.11)** | **< 0.01** |
| Difficulty sleeping | 7,852 | 14 | 1.49 (0.85, 2.62) | 0.17 |
| Felt more tired than normal | 8,522 | 20 | 1.96 (1.21, 3.19) | 0.01 |
| **Severe fatigue (e.g. inability to get out of bed)** | **1,894** | **7** | **3.10 (1.43, 6.69)** | **< 0.01** |
| Numbness or tingling somewhere in the body | 3,651 | 8 | 1.83 (0.89, 3.78) | 0.10 |
| Feeling of heaviness in arms or legs | 2,991 | 7 | 1.96 (0.91, 4.23) | 0.09 |
| Achy muscles | 6,541 | 15 | 1.92 (1.11, 3.32) | 0.02 |
| Symptoms not reported in the last week | 72,678 |  | reference |  |

1) See methods for details of symptom reporting.

### Spatial patterns

We detected differences in crude prevalence at the regional scale, highest in London at 0.20% (95% CI, 0.13%, 0.31%) and lowest in the South West at 0.06% (95% CI, 0.03%, 0.12%) (Figure 4A, Table 7 in Supplement). However higher risk in London was no longer evident after adjustment for age, sex, ethnicity and key worker status (Table 1).

**Table 7.** Prevalence by region.

| Region | All participants | SARS-CoV-2 Positive | Prevalence % (95% CI) |
| --- | --- | --- | --- |
| North East | 4,511 | 5 | 0.11% ( 0.05% , 0.26% ) |
| North West | 15,645 | 24 | 0.15% ( 0.10% , 0.23% ) |
| Yorkshire and The Humber | 7,775 | 7 | 0.09% ( 0.04% , 0.19% ) |
| East Midlands | 15,429 | 18 | 0.12% ( 0.07% , 0.18% ) |
| West Midlands | 11,337 | 19 | 0.17% ( 0.11% , 0.26% ) |
| East of England | 17,598 | 18 | 0.10% ( 0.06% , 0.16% ) |
| London | 10,488 | 21 | 0.20% ( 0.13% , 0.31% ) |
| South East | 27,145 | 41 | 0.15% ( 0.11% , 0.20% ) |
| South West | 10,692 | 6 | 0.06% ( 0.03% , 0.12% ) |

**Figure 4.**
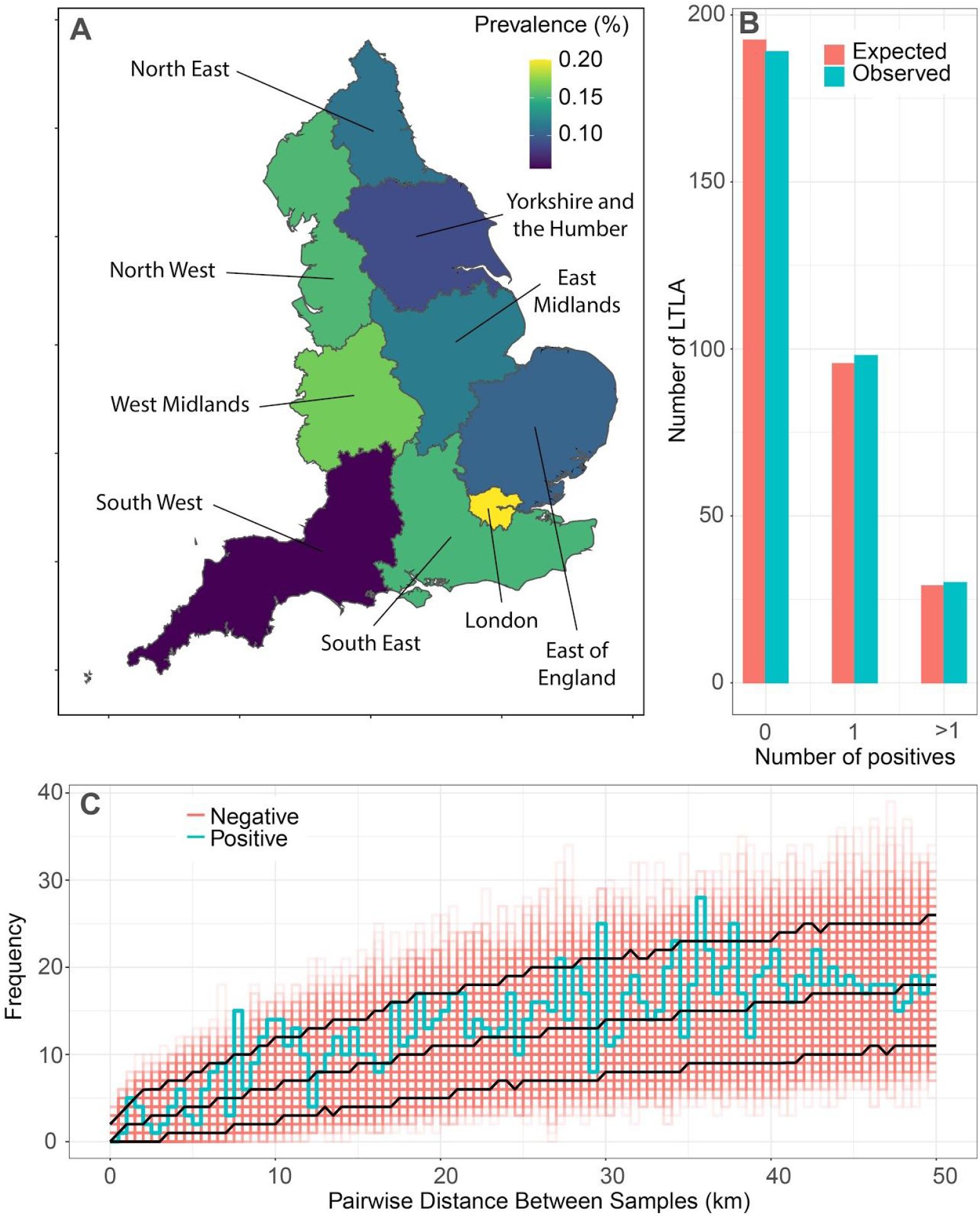
**A** shows regional variation in prevalence of infection. **B** shows observed and expected distribution of swab-positive participants by lower-tier local authority. **C** shows the distribution of pairwise distances between 159 swab-positive participants (blue line) compared with 1000 random redraws of pairwise distances between swab-negative participants (red lines / grid). Black lines show 10th centile, median and 90th centile of the swab-negative distances.

For smaller geographical units, we did not find evidence of spatial clustering. The distribution of LTLAs with 0, 1, and 2 or more cases was consistent with equal prevalence in every LTLA (Figure 4B). Further, the distributions of pairwise distances between the home locations of people testing positive and those testing negative were similar (Figure 4C).

### Sensitivity analysis

We estimated prevalence and decay rate for different Ct values for samples from the commercial laboratory (appendix supplementary methods). The overall unadjusted prevalence of swab positivity decreased from 0.13% to 0.11% (0.095%, 0.13%) for a Ct threshold of 35 and increased to 0.15% (0.13%, 0.18%) for a Ct threshold of 38. The estimated rate of decay of prevalence of 0.081 was similar at 0.083 (0.050, 0.12) per day for a Ct threshold of 35 and reduced to 0.073 (0.044, 0.10) for a Ct threshold of 38.

## Discussion

In this large, nationally representative survey of SARS-CoV-2 infection in England, prevalence was approximately 1 in 1000 and decreasing at the end of the initial lockdown period in May 2020. These observations are in contrast with high model-estimated prevalence at the peak of the epidemic^11^, but are consistent with a recent household-based longitudinal community study^12^. Our results confirm the efficacy throughout lockdown of measures to contain the spread of SARS-CoV-2, with an estimate of R below 0.6 during May.

Although we saw no differences in viral prevalence by sex, we did find age differences with highest rates among young adults (ages 18-24 years) and lowest rates among older people (ages 65+). These patterns were not fully explained by adjustment for sex, key worker status, ethnicity or region, suggesting that differences in social contact behaviour across ages were important. Young adults appear to have maintained higher levels of social contact than other age groups during the lockdown period in England, while older age groups may have effectively shielded^13^. Children and young people aged 5 to 17 yrs had similar rates of infection to adults aged 25 to 44 yrs, indicating that children are similarly susceptible to being infected with SARS-CoV-2^14^. The higher rates of infection in people of Asian (mainly South Asian) ethnicity suggest that some of the increased burden of disease in minority ethnic populations may be due to increased rates of infection^15^.

We found that key workers – and especially care home and health care workers – had markedly increased odds of infection, up to 7-8 fold higher than non-key workers. Thus, care homes and hospitals, even during the period of our study at the end of lockdown, remained an important source of infection. In addition, we found nearly 25-fold greater odds of infection for those individuals who reported coming into contact with a confirmed case of COVID-19, with much of the excess risk experienced by care home and health care workers being explained by their having reported contact with COVID-19 cases. Assuming that many of these contacts were work-related, these results suggest a continuing need for improved infection control in these settings.

We found that a high proportion of swab-positive participants did not report symptoms during the week prior to swabbing, nearly 70%. Current “test and trace” efforts in the UK are focused on care home, health care settings, and symptomatic individuals so will not be triggered by non-symptomatic people. On a given day, they comprise the bulk of infections in the community and an ongoing source of transmission. Therefore, social distancing needs to remain an important component of infection control measures.

We found apparent regional differences in rates, with the highest crude prevalence observed in London, which was a focus of transmission early in the UK epidemic.This may reflect its role as a major transport, business and tourism hub, similar to other global cities such as New York^16^. However, the higher rates in London appear to have been confounded somewhat by other factors including ethnicity and health care worker status. We found no evidence for geographical clustering at a sub-regional or local scale, suggesting that the epidemic in England at this stage was still showing generalized transmission and had not broken down into disconnected sub-epidemics.

Our study has limitations. Although we selected a representative sample of the population from the lists of patients registered with a general practitioner in England (covering almost the entire population) our response rate overall was 31% which may have affected our estimates of prevalence.

Our study involved the use of self-administered nose and throat swabs to obtain evidence of infection. A number of factors need to be considered when interpreting swab-positivity prevalence. First, viral RNA may have been present but not detected during the process of swabbing, transport and analysis. Estimates of diagnostic sensitivity are in the region of 70%^17–19^. Therefore, our estimate of prevalence of swab positivity suggests that up to 115,000 (95% CI 99,000, 134,000) people in England may have been infected with SARS-CoV-2 virus on any given day. Second, viral RNA may persist in the nose and mouth after viable virus is no longer present^20^ and therefore not be indicative of active infection.

Third, it is possible that self-swabbing efficacy may vary according to age and other demographic characteristics. Fourth, we had evidence to suggest that there was differential reporting of a positive result across laboratories with the commercial laboratory initially reporting a higher positivity rate than the PHE laboratories. However, we mitigated the potential impact of this difference through a calibration exercise resulting in similar swab-positivity rates between laboratories. Also, our key findings were robust to different thresholds of RT-PCR Ct-value suggesting that our results were not unduly influenced by Ct threshold.

In conclusion, we suggest nationally-representative population-based surveys of SARS-CoV-2 infection may greatly improve situational awareness. An important feature of the data presented here is that they are independent of service-oriented testing processes, the representativeness of which varies substantially over time and space. Repeated rounds of studies similar to that reported here will enable continued monitoring of key epidemic properties, including R estimates at regional and local levels, to guide locally-optimized interventions.

## Data Availability

The original datasets generated or analysed, or both, during this study are not publicly available because of governance restrictions and the identifiable nature of the data.

## Contributors

SR and PE conceptualised and designed the study and drafted the manuscript. SR, KECA, OE, BJ and CEW undertook the data analysis. PJD, DA and CAD provided statistical advice. GC, WB, HW, CA and AD provided study oversight. GT provided oversight of the laboratory calibration study. AD and PE obtained funding. SR, KECA, OE, BJ, CEW, CA, PJD, DA, CAD, GC, WB, HW, GT, AD and PE critically reviewed the manuscript. All authors read and approved the final version of the manuscript. PE is the guarantor for this paper. The corresponding author attests that all listed authors meet authorship criteria and that no others meeting the criteria have been omitted, had full access to all the data in the study, and had final responsibility for the decision to submit for publication.

## Acknowledgements

We thank key collaborators on this work -- Ipsos MORI: Kelly Beaver, Sam Clemens, Gary Welch, Nick Gilby and Kelly Ward; Institute of Global Health Innovation at Imperial College: Gianluca Fontana, Dr Hutan Ashrafian, Sutha Satkunarajah and Lenny Naar; MRC Centre for Environment and Health, Imperial College London: Daniela Fecht; North West London Pathology and Public Health England for help in calibration of the laboratory analyses; NHS Digital for access to the NHS register; and the Office for Life Sciences and Department of Health and Social Care for logistic support. For comments on drafts of this report, we thank: Michelle Heys, Julia Gog, Adam Kucharski and Christophe Fraser.

## References

1. Coronavirus resource center. Coronavirus resource center. https://coronavirus.jhu.edu/.

2. UK government. Coronavirus (COVID-19) in the UK. Coronavirus (COVID-19) cases in the UK. https://coronavirus.data.gov.uk/. Accessed June 13, 2020.

3. Houlihan C, Vora N, Byrne T, et al. SARS-CoV-2 virus and antibodies in front-line Health Care Workers in an acute hospital in London: preliminary results from a longitudinal study. Infectious Diseases (except HIV/AIDS). June 2020. doi: 10.1101/2020.06.08.20120584

4. Shields AM, Faustini SE, Perez-Toledo M, et al. SARS-CoV-2 seroconversion in health care workers. medRxiv. 2020. https://www.medrxiv.org/content/10.1101/2020.05.18.20105197v1.

5. Graham NSN, Junghans C, Downes R, et al. SARS-CoV-2 infection, clinical features and outcome of COVID-19 in United Kingdom nursing homes. Public and Global Health. May 2020. doi: 10.1101/2020.05.19.20105460

6. McMichael TM, Currie DW, Clark S, et al. Epidemiology of Covid-19 in a Long-Term Care Facility in King County, Washington. N Engl J Med. 2020;382(21):2005-2011. doi: 10.1056/NEJMoa2005412

7. Gandhi M, Yokoe DS, Havlir DV. Asymptomatic Transmission, the Achilles’ Heel of Current Strategies to Control Covid-19. N Engl J Med. 2020;382(22):2158–2160. doi: 10.1056/NEJMe2009758

8. Wearing HJ, Rohani P, Keeling MJ. Appropriate models for the management of infectious diseases. PLoS Med. 2005;2(7):e174. doi: 10.1371/journal.pmed.0020174

9. R Core Team. R: A Language and Environment for Statistical Computing. Vienna, Austria; 2019. https://www.r-project.org/.

10. Wood SN. Generalized Additive Models: An Introduction with R, Second Edition. CRC Press; 2017. https://play.google.com/store/books/details?id=HL-PDwAAQBAJ.

11. Flaxman S, Mishra S, Gandy A, et al. Estimating the effects of non-pharmaceutical interventions on COVID-19 in Europe. Nature. June 2020. doi: 10.1038/s41586-020-2405-7

12. Pouwels KB, House T, Robotham JV, et al. Community prevalence of SARS-CoV-2 in England: Results from the ONS Coronavirus Infection Survey Pilot. doi: 10.1101/2020.07.06.20147348

13. Atchison CJ, Bowman L, Vrinten C, et al. Perceptions and behavioural responses of the general public during the COVID-19 pandemic: A cross-sectional survey of UK Adults. Public and Global Health. April 2020. doi: 10.1101/2020.04.01.20050039

14. Luo L, Liu D, Liao X-L, et al. Modes of contact and risk of transmission in COVID-19 among close contacts. medRxiv. March 2020:2020.03.24.20042606. doi: 10.1101/2020.03.24.20042606

15. Public Health England. Disparities in the Risk and Outcomes of COVID-19 .; 2020.

16. Richardson S, Hirsch JS, Narasimhan M, et al. Presenting Characteristics, Comorbidities, and Outcomes Among 5700 Patients Hospitalized With COVID-19 in the New York City Area. JAMA. April 2020. doi: 10.1001/jama.2020.6775

17. Jiang G, Ren X, Liu Y, et al. Application and optimization of RT-PCR in diagnosis of SARS-CoV-2 infection. Infectious Diseases (except HIV/AIDS). February 2020. doi: 10.1101/2020.02.25.20027755

18. Lo IL, Lio CF, Cheong HH, et al. Evaluation of SARS-CoV-2 RNA shedding in clinical specimens and clinical characteristics of 10 patients with COVID-19 in Macau. Int J Biol Sci. 2020;16(10):1698–1707. doi: 10.7150/ijbs.45357

19. Woloshin S, Patel N, Kesselheim AS. False Negative Tests for SARS-CoV-2 Infection - Challenges and Implications. N Engl J Med. June 2020. doi: 10.1056/NEJMp2015897

20. Guo W-L, Jiang Q, Ye F, et al. Effect of throat washings on detection of 2019 novel coronavirus. Clin Infect Dis. April 2020. doi: 10.1093/cid/ciaa416

